# Exploring non-linear associations of maternal pre-pregnant body mass index with risk of stillbirth, infant and neonatal mortality in over 21 million US births

**DOI:** 10.1101/2023.04.12.23288470

**Authors:** Hannah V Thornton, Rosie P Cornish, Deborah A Lawlor

## Abstract

**Background:** Higher maternal pre-pregnancy body mass index (BMI) has been associated with higher risk of stillbirth, infant and neonatal mortality. Few studies have explored associations of underweight, with those that have varying in their conclusions. Our aim was to examine the risk of stillbirth, infant and neonatal mortality across the pre-pregnancy BMI distribution and establish a likely healthy BMI range.

**Methods:** We used publicly available birth, infant death and fetal death datasets from the US National Center for Health Statistics National Vital Statistics System, 2014–2020. Fractional polynomial multivariable logistic regression was used to examine the nature of associations between maternal pre-pregnant BMI and stillbirth (birth with no signs of life at ≥24 weeks), infant mortality (death of a live born baby aged <365 days) and neonatal mortality (death of a live born baby aged <28 days).

**Findings:** There were 56,376/21,437,556 (0.26%) stillbirths, 108,413/24,742,273 (0.44%) infant deaths and 66,801/24,742,273 (0.27%) neonatal deaths among complete cases. Mean BMI was 27.0 kg/m^2^. We found non-linear associations between pre-pregnant BMI and all three outcomes - risk was elevated at both low and high BMIs although, for stillbirth, the increased risk at low BMI was less marked than for infant and neonatal mortality. The lowest risk was at a BMI of 21 kg/m^2^ for infant and neonatal mortality and, for stillbirth, at 18 kg/m^2^.

**Interpretation:** Public health messaging for preconception and postnatal care should focus on healthy weight to maximise maternal and child health, and not focus solely on maternal overweight or obesity.

## Introduction

Stillbirth (defined as birth with no signs of live between 20 and 28 weeks of gestation, varying between populations and institutions and hence studies), infant mortality (death of a live born baby aged <365 days) and neonatal mortality (death of a live born baby aged <28 days, a subgroup of infant death) are devastating events with severe repercussions for families and healthcare services. Globally, an estimated 2.6 million third trimester (i.e. ≥ 28 weeks) stillbirths occurred in 2015 and, in 2021 over 3.7 million children died before the age of 1; the majority of these deaths occurred in low- and middle-income countries [1, 2]

Internationally, obesity has almost tripled since 1975 and the most recent WHO data shows that an estimated 25% of adults were obese in 2020 [3]. The prevalence of underweight is slowly falling, yet an estimated 9.4% of all females were underweight in 2016 [4]. Whilst higher maternal BMI is consistently associated with a higher risk of stillbirth, neonatal mortality and infant mortality, there is less certainty over the associations of underweight with these outcomes [1, 5] There is an established higher risk of fetal growth restriction and low birth weight in underweight women, and a strong relationship between fetal growth restriction and stillbirth and infant mortality [6]. One might therefore expect an increased risk of stillbirth, neonatal and infant mortality in underweight women.

Of the five previous systematic reviews [5, 7-10] and four subsequent primary studies [11-14] on the relationship of pre-pregnancy BMI on stillbirth, infant or neonatal mortality, all reported results in BMI categories, with three (including two of the reviews) not examining the association of underweight with these outcomes (see research in context panel). These nine studies all reported analyses supporting an increased risk of all three outcomes among women who were overweight or obese, with less consistency regarding underweight. Two of the reviews used information from the BMI categories to estimate a relative risk for a unit increase in BMI and to examine non-linear associations, one with infant and neonatal mortality [10], one with all three outcomes [5]. Both found evidence for non-linearities but, again, these were not consistent, with one indicating a decreasing risk of infant mortality with lower BMIs [10] but the other finding a very slight increase below a BMI of around 20 kg/m^2^ but a decreasing risk of stillbirth with lower BMIs.

Understanding the relationships between maternal underweight, as well as overweight and obesity, and stillbirth, neonatal and infant mortality might enable us to identify a “safe” target weight for women considering pregnancy; this has not yet been identified [5]. This could help parents and healthcare providers make informed decisions about antenatal monitoring, delivery and infant care for mothers whose weight (whether under- or overweight) might identify increased risk of stillbirth and infant mortality.

The aim of this study was to explore whether there were non-linear associations of maternal pre-pregnant BMI with stillbirth, neonatal and infant mortality in order to identify a “healthy” BMI for these outcomes. We add to existing literature by analysing the largest population to date, over 21 million recorded births in the United States (US) between 2014 and 2020, providing sufficient sample size to investigate the risk of these relatively rare outcomes across the whole maternal BMI range.

## Methods

### Dataset

This study used publicly available US National Center for Health Statistics (NHCS) National Vital Statistics System datasets, which contain details of all births registered in the USA [15]. For stillbirths we used the Birth and Fetal Death Data Files from 2014 to 2019; for infant, including neonatal deaths, we used the Period/Cohort Linked Birth-Infant Death Datafiles from 2014 to 2020 [15]. These timeframes were selected because BMI was included in the fetal deaths data from 2014 onwards and, at the time of analysis, fetal death data were available up to and including 2019 and infant death data up to and including 2020. Each record in these datasets relates to a live birth or fetal death, rather than a pregnancy and there are no pregnancy or person-level identifiers in the dataset. To identify pregnancies, we matched multiple births occurring close in time in which birth and maternal characteristics were the same.

### Ethics

These analyses used publicly available data with no identifying information; no ethical approval was required for this study. We have adhered to the Vital Statistics Data User Agreement in the use of these data. The NHCS are bound by federal regulations to ensure data confidentiality.

### Exposure

Maternal pre-pregnancy height and weight were self-reported by the women at the time of birth via the following questions: What is your height? What was your pre-pregnancy weight? That is, your weight immediately before you became pregnant with this child? BMI was calculated from these. For the descriptive and secondary analysis, World Health Organisation (WHO) BMI categories were used, including categories of underweight (severe underweight: <16 kg/m^2^; moderate underweight: 16-16.9 kg/m^2^; mild underweight: 17-18.49 kg/m^2^; normal: 18.5-24.9 kg/m^2^; overweight 25-29.9 kg/m^2^; obesity classes I, II, and III: 35-34.9 kg/m^2^, 35-39.9 kg/m^2^ and ≥ 40 kg/m^2^.

### Outcomes

Gestational age at which a fetal death is classified as a stillbirth varies internationally. The WHO define stillbirth as a baby born with no signs of life at ≥28 weeks’ gestation. In many European countries a threshold of gestational age of ≥22 weeks is used; 24 weeks is used in the United Kingdom and 20 weeks in the US [16]. To enable comparison, we report results using three different definitions. For the main results we used ≥24 weeks’ gestation, with results using ≥20 and ≥28 weeks also reported. WHO definitions were used for infant mortality (death of a live born baby aged <365 days) and neonatal mortality (death of a live born baby aged <28 days).

### Potential confounders

The following variables were considered to be potential confounders, based on their known or plausible causal effect on maternal BMI and stillbirth, and infant, including neonatal, mortality: maternal age at birth (categorised as: <20, 20-24, 25-29, 30-34, 35-39, 40+ years); maternal education (<high school, high school, college, no degree, degree/higher); maternal ethnic group (White, Black, American Indian/Alaskan Native, Asian, Native Hawaiian/Pacific Islander, Mixed race); smoking status (non-smoker, stopped in early pregnancy, smoked throughout pregnancy); any other live birth in the previous 12 months (yes, no); parity (0, 1, 2, 3, ≥4); multiple birth (yes/no).

### Statistical analysis

Characteristics of the analysis sample were compared with the whole sample to identify any patterns of missingness. Since the proportion of missing data was relatively small (just over 90% had complete data), a complete case analysis was used. For the primary analysis, multivariable logistic regression using fractional polynomials [17] with up to two powers were used to explore potential non-linear relationships between pre-pregnant BMI and each of the three outcomes. Robust standard errors were used to take account of non-independence of outcomes of multiple births. We describe the overall shape of associations from these fractional polynomial regression analyses and, where there was evidence of a pre-pregnant BMI with lowest risk, we estimated that BMI value using differentiation and used bootstrapping to obtain its confidence interval. In a secondary analysis, we used BMI categories in a logistic regression.

## Results

The stillbirth dataset (24-week definition) contained 23,444,074 birth records of which 21,437,556 (91%) had no missing data (Figure 1); the infant mortality dataset contained 26,981,729 birth records of which 24,472,273 (91%) had no missing data (Figure 2). Among the complete case (i.e. no missing data) analysis samples, there were 56,376 (0.26%) stillbirths at or after 24 completed weeks, 108,413 (0.44%) infant deaths, and 66,801 (0.27%) neonatal deaths. Stillbirth proportions were 0.41% and 0.20% with gestational age thresholds of 20 and 28 weeks, respectively. All three outcomes were slightly less common in the complete case sample compared to the whole sample (0.26% vs 0.33% for stillbirth; 0.27% vs 0.38% for neonatal mortality and 0.44% vs 0.56% for infant mortality), with distributions of all other variables being similar (Supplementary Tables 1 and 2).

**Figure 1:**
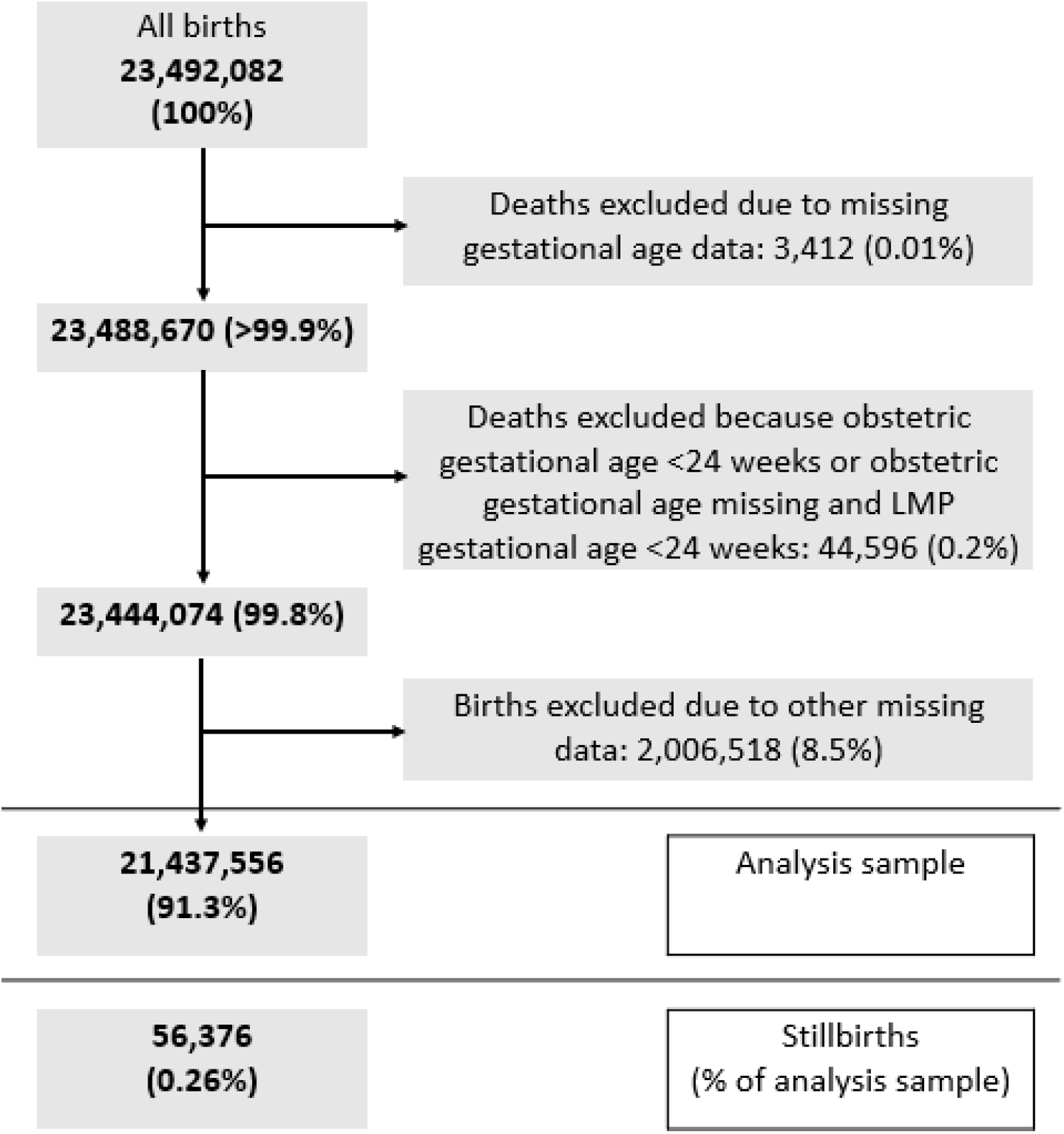
Stillbirth data inclusion flowchart (stillbirths after 24 weeks gestation)

**Figure 2:**
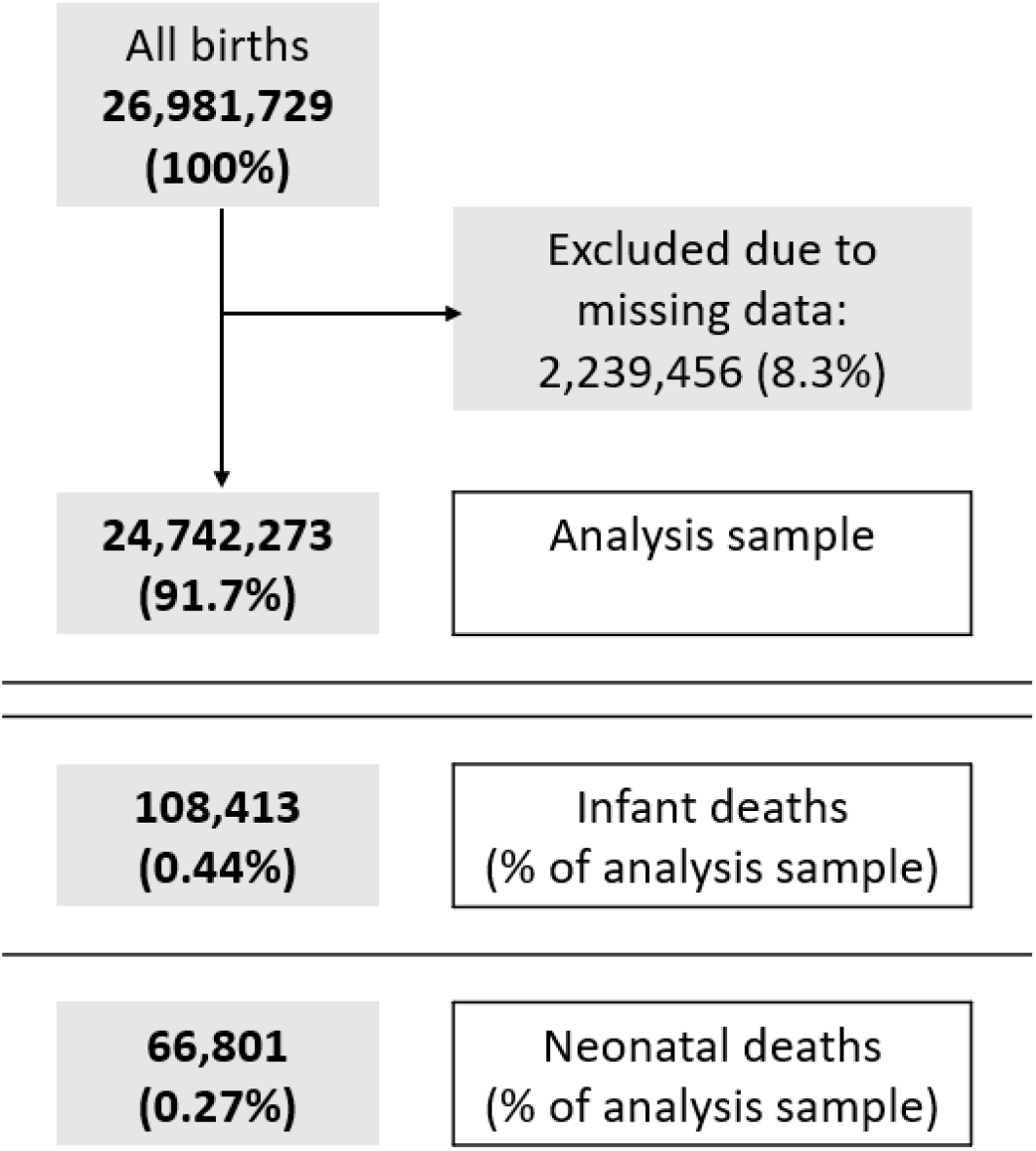
Infant mortality data inclusion flowchart

Details of the fractional polynomial models are given in the supplementary text. Figures 3 to 5 give predicted risks (adjusted for confounders) across the BMI range obtained from these models. For all three outcomes there was evidence for a j-shaped curve, with an increased risk at very low maternal pre-pregnant BMIs as well as at higher BMIs. The increased risk among underweight women was more marked for infant and neonatal mortality than for stillbirths. The maternal BMI at which the predicted risk was lowest was 18.1 kg/m^2^ (95% CI: 15.2, 22.4 kg/m^2^) for stillbirth, 21.3 kg/m^2^ (21.1, 21.5 kg/m^2^) for infant mortality, and 20.6 kg/m^2^ (17.7, 24.5 kg/m^2^) for neonatal mortality.

**Figure 3:**
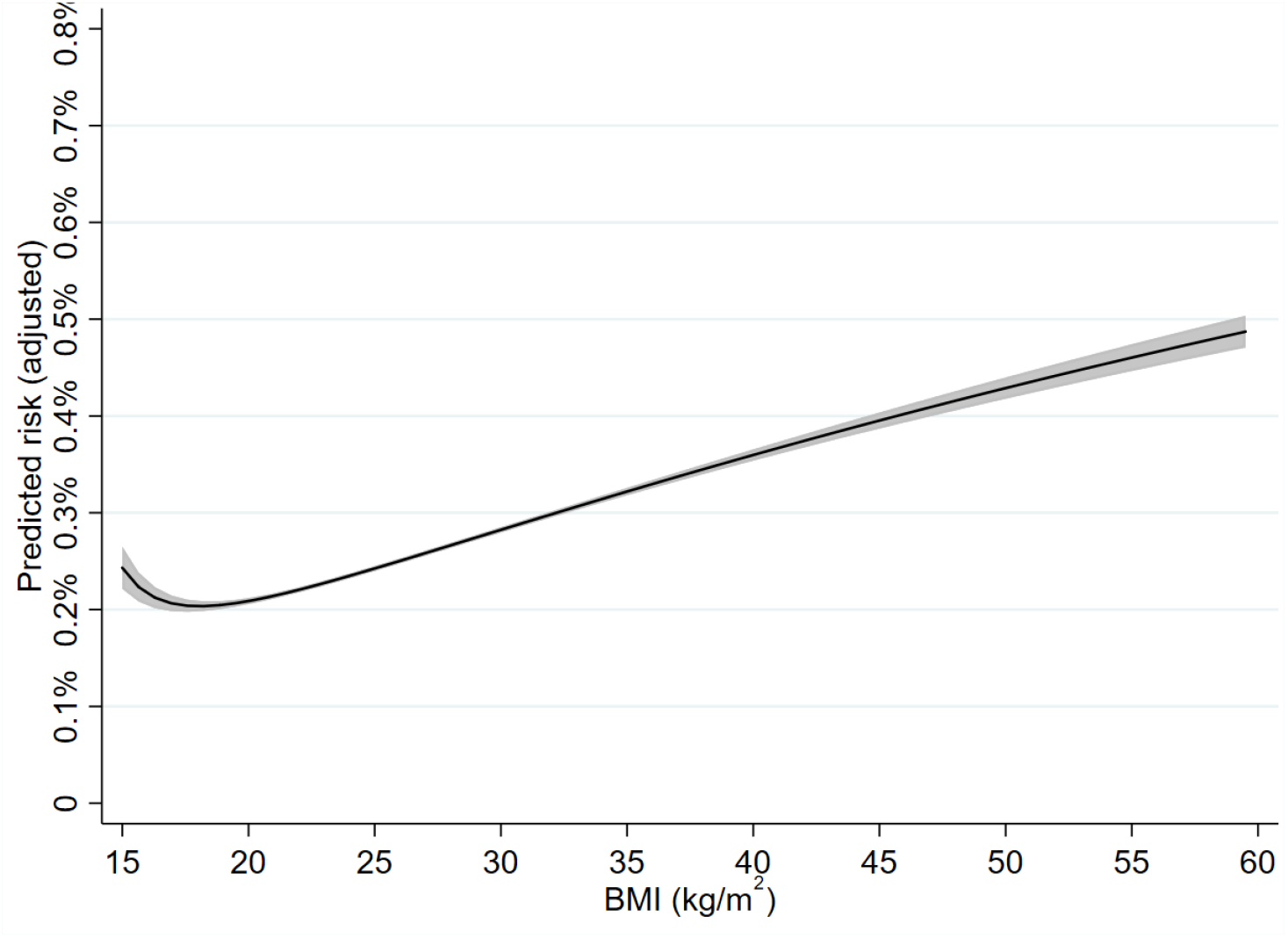
Adjusted risk of stillbirth by maternal pre-pregnant BMI (kg/m^2^) Footnote: Among 21.4 million US women with deliveries between 2014 and 2019 (inclusive)

**Figure 4:**
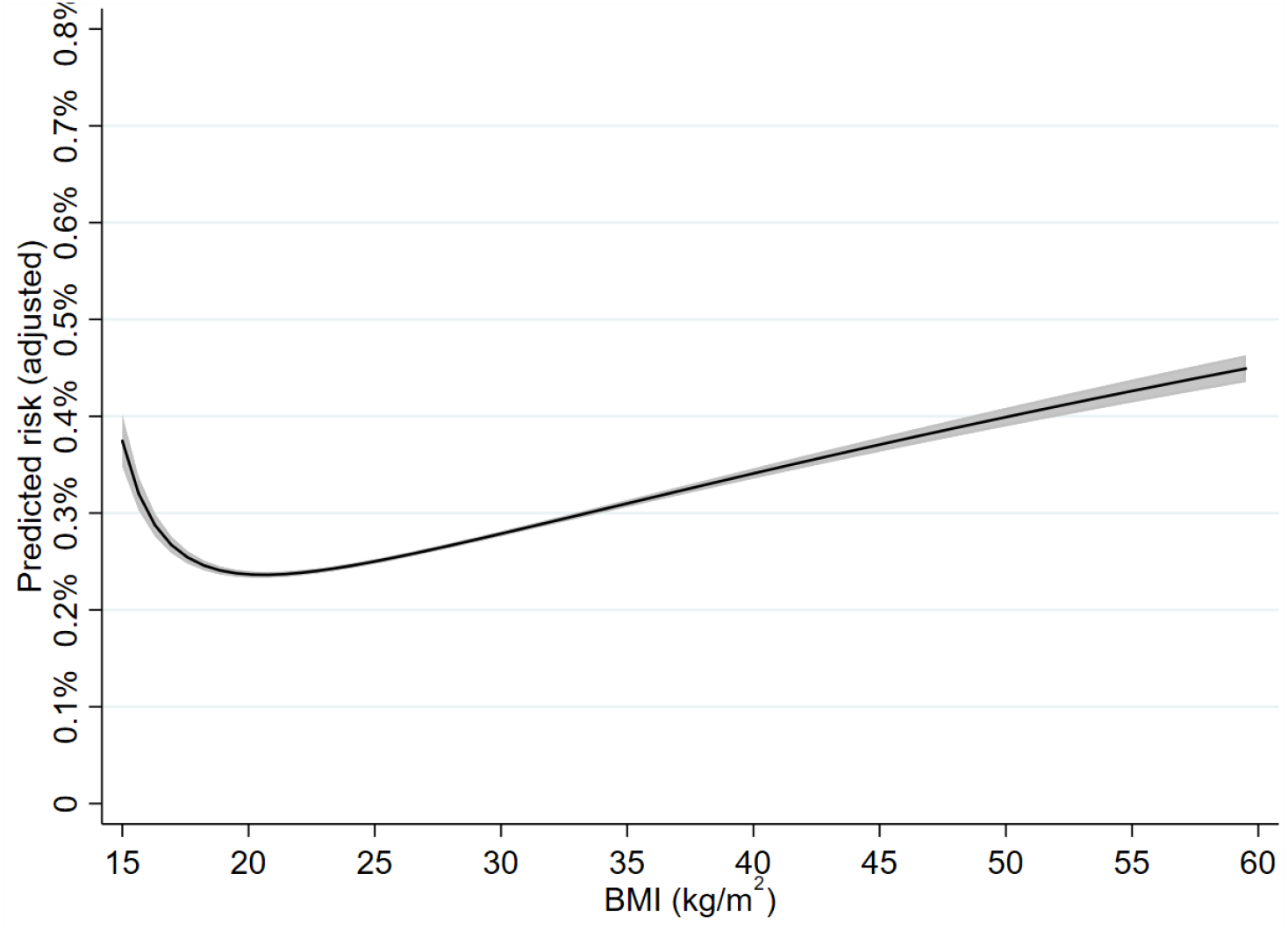
Adjusted risk of neonatal mortality by maternal pre-pregnant BMI (kg/m^2^) Footnote: Among 24.7 million US women with deliveries between 2014 and 2020 (inclusive)

**Figure 5:**
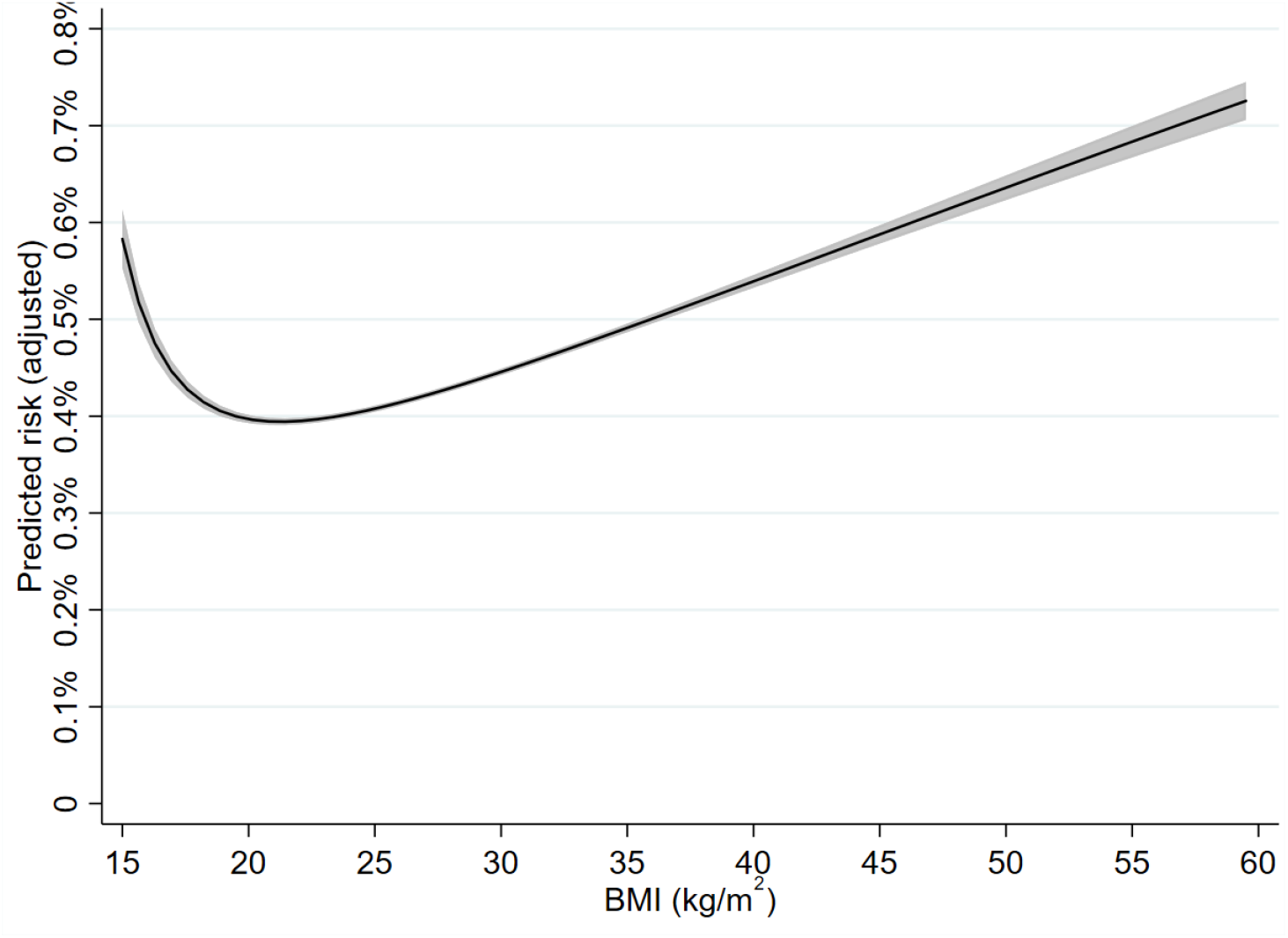
Adjusted risk of infant mortality by maternal pre-pregnant BMI (kg/m^2^) Footnote: Among 24.7 million US women with deliveries between 2014 and 2020 (inclusive)

The overall pattern of association for stillbirth remained similar when thresholds of 20 and 28 completed weeks of gestation were used, although predicted risks of stillbirth were higher when a 20-week threshold was used (compared to 24 weeks) and lower when a 28-week threshold was used. In addition, the increased risk for low BMIs was more marked with a 20-week threshold and the increase with higher BMIs was also more marked (Supplementary Figure 1).

Unadjusted risks of stillbirth, infant mortality and neonatal mortality using BMI categories are given in Supplementary Tables 3 and 4 and adjusted odds ratios given in Supplementary Table 5; adjusted odds ratios for stillbirths using the 20-week and 28-week threshold are given in Supplementary Table 6. The patterns broadly matched those shown in Figures 3 to 5, with an increased risk of infant and neonatal mortality across all underweight categories whereas, for stillbirth, the risk compared to normal weight women is only elevated among those who are severely underweight; for all three outcomes risk was higher in women who were overweight and those in all categories of obesity.

## Discussion

In this study of over 21 million births, we have provided evidence for j-shaped associations between maternal pre-pregnant body mass index and stillbirth, infant mortality and neonatal mortality, with an increased risk among both underweight and overweight/obese women. This increased risk for underweight women was more marked for infant and neonatal mortality and only very slight for stillbirth. The lowest predicted risk occurred at a BMI of just over and just under 21 kg/m^2^ for infant and neonatal mortality (respectively) and at a BMI of just over 18 kg/m^2^ for stillbirth. Whilst the obesity epidemic has shifted some aspects of preconceptual advice and antenatal care to a focus avoiding overweight and obesity in women of reproductive age and during pregnancy [18], our results demonstrate that that in contemporary high-income populations who have experienced several decades of the obesity epidemic, stillbirth, neonatal and infant mortality risk are higher in women with both higher and lower BMI. Taken together, the confidence intervals from our study suggest that women with pre-pregnancy BMI below 15.2 kg/m^2^ or above 24.5 kg/m^2^ are at increased risk of all three outcomes, with values around 20-22kg/m^2^ likely associated with lowest risk of all three.

Previous studies have consistently found an increased risk of these outcomes with overweight and obesity categories but the results for underweight have been less consistent. In the most recent and largest meta-analysis of 13.5 million births, underweight was found to be associated with a decreased risk of infant mortality [10]. However, this result was dominated by one large study (receiving 83% of the weight) [10] in which the odds ratios were adjusted for gestational age and small for gestational age. These factors are very likely to be on the causal pathway between maternal BMI and infant mortality, thus meaning that this is not testing the effect of underweight, much of which is likely due to the impact of maternal BMI on preterm and small for gestational age [19]. To our knowledge, only two previous studies have analysed the association between pre-pregnant BMI and these outcomes using maternal BMI as a continuous variable. One examined the association with fetal or infant death as a combined outcome and found evidence for a V-shaped relationship with a minimum at a BMI of 23 kg/m^2^ [20], but numbers in this study were relatively small (N = 40,932), and hence results imprecise. The other investigated whether there was evidence for a quadratic relationship between maternal BMI and stillbirth in different subgroups of ethnicity, parity and gestational age but underweight women were excluded due to small numbers [21]. We have added to previous work by determining the pattern of association across the whole maternal distribution and deriving BMI levels at which risk for all three of stillbirth, infant death and its subgroup of neonatal death are lowest.

Pre-pregnancy higher BMI is associated with an increased risk of pregnancy complications including hypertensive disorders of pregnancy and gestational diabetes, which are in turn associated with an increased risk of stillbirth and neonatal mortality [22, 23], likely via impaired placental function and consequent fetal growth restriction and/or preterm birth [1, 24] or – in the case of gestational diabetes - complications arising as a result of fetal macrosomia [23]. Lower maternal BMI is associated with an increased risk of small for gestational age and low birthweight [8], which are strongly associated with stillbirth and infant death [25]. Whilst recent integration of evidence from conventional multivariable regression, Mendelian randomization and a negative control analyses support a causal effect of higher maternal BMI on pre-eclampsia, gestational hypertension, and macrosomia / large for gestational age, and a lower risk of small for gestational age, even linear effects on stillbirth were too imprecise to make robust conclusions [26]. Preterm birth is also a risk factor for infant mortality and evidence suggests non-linear associations of maternal pre-pregnancy BMI with preterm birth, with higher BMI largely contributing to increased risk of medically indicated BMI, and lower BMI to spontaneous preterm birth [27, 28].

Key strengths of this study include the large sample size and the fact that we have explored associations with BMI as a continuous variable. Although the distribution of most characteristics was similar among the whole sample and complete cases, data were less complete for stillbirths and births which resulted in infant deaths. However, a complete case logistic regression will give unbiased estimates of the exposure odds ratio when missingness only depends on the outcome [29] and the proportion of missing data was relatively small (<10%). Thus, we do not believe that missing data will have had an important impact on the results. We adjusted for all plausible confounding factors but cannot assume that increased risk at higher or lower BMI causes these outcomes. It is not possible to randomize women to different BMI levels and whilst our recent Mendelian randomization analyses (discussed above) provides evidence of causal effects on outcomes such as hypertensive disorders of pregnancy and gestational diabetes that might mediate maternal BMI effects on stillbirth, infant and neonatal mortality, non-linear Mendelian randomization for these rare outcomes is currently unfeasible as no study or collaboration of studies would have sufficiently large numbers with genomic data and these outcomes. Finally, our analysis is of women resident and pregnant in the US between 2014 and 2020 and may not generalise to other populations, particularly those at a different stage of the obesity epidemic. If these results were replicated in different populations at different stages of the obesity epidemic and with different ethnic and socio-economic distributions that might increase our confidence in them being causal.

In conclusion, in this large contemporary high-income population we show that risk of stillbirth, infant mortality and neonatal mortality is increased at lower and higher maternal BMI, with pre-pregnancy BMI values within the range 20 to 22 kg/m^2^ likely to minimise risk for all three of these rare but devastating outcomes. Non-linear associations of maternal BMI are evident for birth weight, with higher BMI relating to large-for-gestational age and lower BMI linked to increased risk of small for gestational age, and for preterm birth [30]. Taken together with results we present here, this emerging body of research highlights the need for public health advice to women of reproductive age to recognise that lower, as well as higher BMI is associated with adverse maternal and offspring outcomes. Future research should aim to understand the factors and mechanisms which underlie these associations, and refine the range of BMI levels that optimise all aspects of maternal and offspring perinatal health.

## Data Availability

All data used in this study are available online at

https://www.cdc.gov/nchs/data_access/vitalstatsonline.htm

