## Supplementary Materials for "Exploring non-linear associations of maternal pre-pregnant body mass index with risk of stillbirth, infant and neonatal mortality in over 21 million US births"

A. Supplementary text

**Fractional polynomial models**

To fit the fractional polynomials, BMI was first scaled [scaled BMI = (BMI-10)/5]. This is done because if the values of the variable are too large (or too small), this can generate extreme values with certain powers of this variable (e.g. cubic or squared reciprocal powers).

Results

For stillbirth and infant neonatal mortality, the best fitting two-degree fractional polynomial model had powers of -1 and -0.5 - i.e. the reciprocal of (scaled) BMI and one over the square root of (scaled) BMI. For neonatal mortality, the best fitting two-degree model had powers of -0.5 and -0.5 – i.e. one over the square root of (scaled) BMI and the natural logarithm of scaled BMI multiplied by one over the square root of (scaled) BMI. For each outcome, this two-degree model fitted better than the best fitting one-degree model (difference in deviance = 186.6 for infant death, 147.9 for neonatal death and 41.9 for stillbirth; p<0.001 in all cases).

B. Supplementary tables and figures

Supplementary Table 1: Characteristics of whole sample compared to analysis sample: stillbirth dataset (24-week definition)

| Characteristic | Whole sample N=24,444,074[1] | Analysis sample N=21,437,556 |
|---|---|---|
| **Stillbirth** | 77,206 (0.33%) | 56,376 (0.26%) |
| **Maternal pre-pregnant BMI** | | |
| <18.5 | 770,923 (3%) | 732,480 (3%) |
| 18.5-24.9 | 10,043,599 (45%) | 9,556,159 (45%) |
| 25-29.9 | 5,869,645 (26%) | 5,564,679 (26%) |
| 30-34.9 | 3,133,544 (14%) | 2,970,761 (14%) |
| 35-39.9 | 1,607,409 (7%) | 1,527,455 (7%) |
| 40+ | 1,141,365 (5%) | 1,086,022 (5%) |
| **Maternal age** | | |
| <20 | 1,254,839 (5%) | 1,177,634 (5%) |
| 20-24 | 4,757,383 (20%) | 4,419,657 (21%) |
| 25-29 | 6,786,386 (29%) | 6,242,664 (29%) |
| 30-34 | 6,596,395 (28%) | 5,996,334 (28%) |
| 35-39 | 3,299,323 (14%) | 2,947,748 (14%) |
| 40+ | 749,090 (3%) | 653,519 (3%) |
| **Maternal education** | | |
| < High school | 3,126,324 (14%) | 2,856,954 (13%) |
| High school | 5,840,933 (25%) | 5,440,332 (25%) |
| College, no degree | 6,615,258 (29%) | 6,224,387 (29%) |
| Degree/higher | 7,335,731 (32%) | 6,915,583 (32%) |
| **Maternal ethnicity** | | |
| White | 17,195,798 (74%) | 15,946,056 (74%) |
| Black | 3,672,664 (16%) | 3,293,919 (15%) |
| American Indian / Alaskan Native | 225,815 (1%) | 206,637 (1%) |
| Asian | 1,549,778 (7%) | 1,392,996 (6%) |
| Native Hawaiian / Pacific Islander | 75,027 (0.3%) | 61,481 (0.3%) |
| Mixed race | 582,002 (3%) | 536,467 (3%) |
| **Smoking status** | | |
| Non smoker | 20,875,335 (91%) | 19,415,049 (91%) |
| Stopped early pregnancy | 762,749 (3%) | 724,747 (3%) |
| Smoked throughout pregnancy | 1,400,197 (6%) | 1,297,760 (6%) |
| **Parity** | | |
| 0 | 8,958,343 (38%) | 8,498,348 (40%) |
| 1 | 7,480,853(32%) | 6,747,071 (31%) |
| 2 | 3,991,852 (17%) | 3,577,897 (17%) |
| 3 | 1,726,503 (7%) | 1,538,021 (7%) |
| 4+ | 1,230,309 (5%) | 1,076,219 (5%) |
| **Other live birth in previous 12 months** (=yes) | 194,229 (1%) | 183,441 (1%) |
| **Multiple birth** (=yes) | 802,951 (3%) | 704,112 (3%) |

1. Denominators vary because not all variables are complete

Supplementary Table 2: Characteristics of whole sample compared to analysis sample: infant and neonatal mortality dataset

| **Characteristic** | **Whole sample N=26,981,729[1]** | **Analysis sample N=24,742,273** |
|---|---|---|
| **Infant death** | 150,794 (0.56%) | 108,413 (0.44%) |
| **Neonatal death** | 102,382 (0.38%) | 66,801 (0.27%) |
| **Maternal pre-pregnant BMI (kg/m$^2$)** | | |
| <18.5 | 869,042 (3%) | 825,736 (3%) |
| 18.5-24.9 | 11,468,689 (44%) | 10,914,191 (44%) |
| 25-29.9 | 6,811,643 (26%) | 6,458,030 (26%) |
| 30-34.9 | 3,665,925 (14%) | 3,473,944 (14%) |
| 35-39.9 | 1,877,712 (7%) | 1,792,484 (7%) |
| 40+ | 1,344,431 (5%) | 1,277,888 (5%) |
| **Maternal age** | | |
| <20 | 1,409,908 (5%) | 1,323,813 (5%) |
| 20-24 | 5,407,170 (20%) | 5,033,014 (21%) |
| 25-29 | 7,790,269 (29%) | 7,185,669 (29%) |
| 30-34 | 7,646,649 (28%) | 6,977,247 (28%) |
| 35-39 | 3,852,173 (14%) | 3,456,007 (14%) |
| 40+ | 875,117 (3%) | 766,523 (3%) |
| **Maternal education** | | |
| < High school | 3,538,527 (13%) | 3,235,634 (13%) |
| High school | 6,764,960 (26%) | 6,307,956 (25%) |
| College, no degree | 7,583,360 (29%) | 7,139,630 (29%) |
| Degree/higher | 8,532,779 (32%) | 8,059,053 (33%) |
| **Maternal ethnicity** | | |
| White | 19,800,230 (74%) | 18,392,837 (74%) |
| Black | 4,238,581 (16%) | 3,807,744 (15%) |
| American Indian / Alaskan Native | 260,078 (1%) | 237,909 (1%) |
| Asian | 1,778,345 (7%) | 1,601,191 (6%) |
| Native Hawaiian / Pacific Islander | 87,389 (0.3%) | 72,247 (0.3%) |
| Mixed race | 682,344 (3%) | 630,345 (3%) |
| **Smoking status** | | |
| Non smoker | 24,108,710 (91%) | 22,483,026 (91%) |
| Stopped early pregnancy | 848,141 (3%) | 806,948 (3%) |
| Smoked throughout pregnancy | 1,564,524 (6%) | 1,452,299 (6%) |
| **Parity** | | |
| 0 | 10,324,497 (38%) | 9,805,730 (40%) |
| 1 | 8,597,601(32%) | 7,784,036 (31%) |
| 2 | 4,588,551 (17%) | 4,126,536 (17%) |
| 3 | 1,989,397 (7%) | 1,777,625 (7%) |
| 4+ | 1,422,122 (5%) | 1,248,346 (5%) |
| **Other live birth in previous 12 months** (=yes) | 225,645 (1%) | 211,948 (1%) |
| **Multiple birth** (=yes) | 913,447 (3%) | 799,316 (3%) |

1. Denominators vary because not all variables are complete

Supplementary Table 3: Numbers and unadjusted risks of stillbirth (24-week threshold, among analysis sample) by maternal pre-pregnant BMI categories

| Maternal pre-pregnant BMI category | N | Number of stillbirths (risk) |
|---|---|---|
| Severe underweight: <16 | 51,361 | 141 (0.27%) |
| Moderate underweight: 16 – 16.9 | 119,781 | 279 (0.23%) |
| Mild underweight: 17 – 18.49 | 561,338 | 1,143 (0.20%) |
| Normal weight: 18.5 – 24.9 | 9,556,159 | 20,161 (0.21%) |
| Overweight: 25 – 29.9 | 5,564,679 | 14,831 (0.27%) |
| Obesity class I: 30 – 34.9 | 2,970,761 | 9,598 (0.32%) |
| Obesity class II: 35 – 39.9 | 1,527,455 | 5,497 (0.36%) |
| Obesity class III: 40+ | 1,086,022 | 4,726 (0.44%) |

Supplementary Table 4: Numbers and unadjusted risks of infant and neonatal mortality (among analysis sample) by maternal pre-pregnant BMI categories

| Maternal pre-pregnant BMI category | N | Number (risk) | |
|---|---|---|---|
| | | Infant mortality | Neonatal mortality |
| Severe underweight: <16 | 58,083 | 382 (0.66%) | 226 (0.39%) |
| Moderate underweight: 16 – 16.9 | 134,728 | 713 (0.53%) | 395 (0.29%) |
| Mild underweight: 17 – 18.49 | 632,925 | 2,895 (0.20%) | 1,688 (0.27%) |
| Normal weight: 18.5 – 24.9 | 10,914,191 | 40,788 (0.37%) | 24,795 (0.23%) |
| Overweight: 25 – 29.9 | 6,458,030 | 26,995 (0.42%) | 16,769 (0.26%) |
| Obesity class I: 30 – 34.9 | 3,473,944 | 17,507 (0.50%) | 10,886 (0.31%) |
| Obesity class II: 35 – 39.9 | 1,792,484 | 10,393 (0.58%) | 6,556 (0.37%) |
| Obesity class III: 40+ | 1,277,888 | 8,740 (0.68%) | 5,486 (0.43%) |

Supplementary Table 5: Adjusted odds ratios (95% CI) for stillbirth, infant mortality and neonatal mortality by maternal pre-pregnant BMI categories

| Maternal pre-pregnant BMI category | Stillbirth (24 weeks) | Infant mortality | Neonatal mortality |
|---|---|---|---|
| Severe underweight: <16 | 1.13 (0.96, 1.34) | 1.37 (1.23,1.52) | 1.43 (1.25, 1.64) |
| Moderate underweight: 16 – 16.9 | 0.99 (0.87, 1.11) | 1.14 (1.06, 1.23) | 1.11 (1.00, 1.23) |
| Mild underweight: 17 – 18.49 | 0.91 (0.86, 0.97) | 1.08 (1.04, 1.12) | 1.08 (1.03 (1.14) |
| Normal weight: 18.5 – 24.9 | 1.00 | 1.00 | 1.00 |
| Overweight: 25 – 29.9 | 1.19 (1.17, 1.22) | 1.05 (1.03, 1.06) | 1.08 (1.06, 1.10) |
| Obesity class I: 30 – 34.9 | 1.39 (1.35, 1.42) | 1.18 (1.16, 1.20) | 1.23 (1.20, 1.26) |
| Obesity class II: 35 – 39.9 | 1.51 (1.46, 1.56) | 1.30 (1.27, 1.33) | 1.37 (1.34, 1.41) |
| Obesity class III: 40+ | 1.76 (1.71, 1.82) | 1.46 (1.42, 1.49) | 1.52 (1.48, 1.57) |

Supplementary Table 6: Adjusted odds ratios (95% CI) for stillbirth using the 20-week and 28-week thresholds

| Maternal pre-pregnant BMI category | Stillbirth (20 weeks) | Stillbirth (28 weeks) |
|---|---|---|
| Severe underweight: <16 | 1.11 (0.96, 1.27) | 1.03 (0.84, 1.26) |
| Moderate underweight: 16 – 16.9 | 1.08 (0.98, 1.19) | 0.95 (0.83, 1.09) |
| Mild underweight: 17 – 18.49 | 0.93 (0.88, 0.98) | 0.89 (0.83, 0.95) |
| Normal weight: 18.5 – 24.9 | 1.00 | 1.00 |
| Overweight: 25 – 29.9 | 1.19 (1.17, 1.21) | 1.20 (1.17, 1.23) |
| Obesity class I: 30 – 34.9 | 1.40 (1.37, 1.43) | 1.39 (1.35, 1.43) |
| Obesity class II: 35 – 39.9 | 1.58 (1.54, 1.62) | 1.54 (1.49, 1.59) |
| Obesity class III: 40+ | 1.80 (1.76, 1.85) | 1.75 (1.69, 1.82) |

Supplementary Figure 1: Predicted risk of stillbirth (adjusted) by maternal pre-pregnant BMI for the different stillbirth definitions (shaded area indicates 95% confidence intervals)

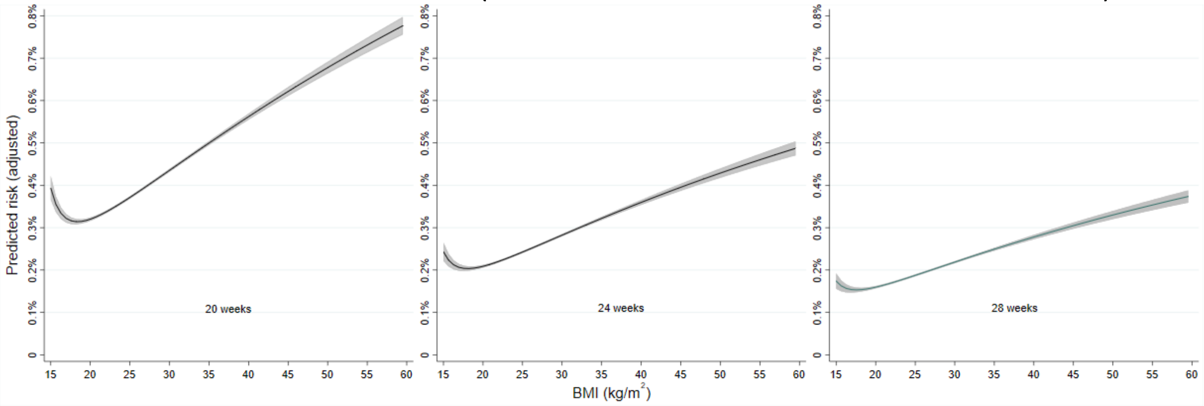
